# Prescribers’ and Pharmacists’ Perception of Adoption and Implementation of Electronic Prescribing and Medication Management System in Public Health Facilities of Dessie city, Northeast Ethiopia

**DOI:** 10.1101/2025.01.23.25321028

**Authors:** Getachew Moges, Naser Ebrahim, Ewunetie Mekashaw Bayked, Teshager Aklilu Yesuf, Solomon Ahmed Mohammed

## Abstract

Medication errors are one of the major causes of preventable adverse drug effects. Errors associated with prescribing and dispensing are prevalent. Electronic prescribing (e-prescribing) offers a solution to mitigate these errors by enhancing legibility, dosing, and remote accessibility. This study aimed to investigate the perceptions of prescribers and pharmacists regarding the adoption and implementation of electronic prescribing (e-prescribing) and electronic medication management (EMM) systems in public health facilities of Dessie City administration, Northeast Ethiopia. A cross-sectional survey was conducted from April 20 to June 25, 2022, in public health facilities of Dessie City administration. A structured self-administered questionnaire was employed to collect data from 201 prescribers and pharmacists. The study was based on the unified theory of acceptance and use of technology model. Data were analyzed using SPSS version 25 and SmartPLS version 4.0. Nearly 34 (72.5%) of the respondents were males. The majority (88.6%) of the participants had a positive perception towards the use of e-prescribing and EMM systems. The mean score for performance expectancy, effort expectancy, and facilitating conditions were 4.29, 4.26, and 4.18, respectively. Perceived facilitating conditions (β = 0.271, p = 0.003), price value (β = 0.239, p = 0.002), performance expectancy (β = 0.237, p = 0.008), and effort expectancy (β = 0.176, p = 0.008) were found to affect the acceptance of e-prescribing and EMM systems among prescribers and pharmacists. Prescribers and pharmacists have a positive perception towards the use of e-prescribing and EMM systems. Perceived facilitating conditions, price value, performance expectancy, and effort expectancy significantly influence acceptance. The effect of perceived credibility on acceptance was fully mediated by performance expectancy.

**Author Summary:** Despite the demonstrated effectiveness of e-prescribing and Electronic Medication Management (EMM) systems in reducing medication errors, their adoption in developing countries has been sluggish. Furthermore, there have been reports of a high rate of failure to realize the anticipated benefits following the implementation of electronic information systems. The acceptance of technology by healthcare professionals is one of the major factors influencing the success or failure of health information system implementation. Most of the studies conducted in Africa regarding the acceptance of technology in healthcare predominantly focused on e-prescribing systems. In Ethiopia, studies on technology acceptance within the healthcare sector are limited, with a few studies focusing on the acceptance of EMM systems. To our search, no study has been conducted to assess physicians’ and/or pharmacists’ perceptions regarding the acceptance of e-prescribing and EMM systems in Ethiopia. Our study aims to assess the acceptance of e-prescribing and EMM systems by prescribers and pharmacists and the factors influencing the acceptance of these systems in public health facilities of Dessie city administration, North East Ethiopia using a modified Unified Theory of Acceptance and Use of Technology (UTAUT) model. The findings of our study are likely to be a great interest to policy makers, researchers, clinicians, and trainees.

## Introduction

Medication errors are any faults in the medication process that may or may not result in patient harm (1). Medication errors are a major public health issue (2). They are one of the major causes of preventable adverse drug effects (3, 4). Of all medication errors reported, those related to prescribing and dispensing are the most common (5).

Traditionally, prescriptions have been handwritten or printed and manually signed by prescribers on paper, subsequently presented by patients to pharmacists for dispensation. However, this method has inherent limitations, including errors arising from prescribers’ poor handwriting and security concerns associated with lost or forged prescriptions (6). However, the use of information and communication technologies in Healthcare can enhance the quality and safety of the medication process (7).

Electronic prescribing (e-prescribing) systems can reduce errors associated with prescribing and dispensing (4, 8–11). (They decrease the likelihood of errors due to illegible handwriting and minimize dosing errors by specifying commonly used doses. Additionally, these systems help prevent adverse drug reactions by identifying potential allergies. Furthermore, e-prescribing enables remote accessibility, promoting efficient communication and coordination among healthcare providers (12, 13).

Electronic medication management systems (EMM) incorporating electronic medicine datasets such as prescribing, administration, pharmacy verification, automated dispensing cabinets, and barcode medication administration, hold the potential to reduce medication errors and improve the quality of the medication process (14). Studies indicated that computerized physician order entry systems contribute to reducing medication errors (4) (15). Furthermore, the integration of these systems has been shown to yield additional reductions in errors (4, 15). (16).

Pharmaceutical operations within health facilities must be optimized to improve the performance of healthcare workers and patient convenience and satisfaction (17). The adoption of a technological system by individuals depends to a considerable extent on perceived usefulness and ease of use (18). The complexities and inconsistencies among e-prescribing products have been linked to their limited utilization (19). Remarkably, only 5% of physicians prescribed electronically in the year 2000 (20). Physicians often hesitate to adopt e-prescribing due to the system not aligning with their routine practice and the loss of interest in technology (21). Moreover, some find these systems time-consuming and a hindrance to the physician-patient relationship (12, 22). Variations in work patterns among pharmacists have been also observed utilizing EMM and those that do not (23).

Despite the demonstrated effectiveness of e-prescribing and EMM systems in reducing medication errors, their adoption in developing countries has been sluggish. Furthermore, there have been reports of a high rate of failure to realize the anticipated benefits following the implementation of electronic information systems (24). The acceptance of technology by healthcare professionals is one of the major factors influencing the success or failure of health information system implementation (25).

The majority of the studies conducted in Africa regarding the acceptance of technology in healthcare predominantly focused on e-prescribing systems (26–32). However, in Ethiopia, studies on technology acceptance within the healthcare sector have been limited, with a few studies focusing on the acceptance of EMM systems (26, 33–36). To the best of the authors’ knowledge, no study has been conducted to assess physicians’ and/or pharmacists’ perceptions regarding the acceptance of e-prescribing and EMM systems in Ethiopia. Therefore, this study aims to assess the acceptance of e-prescribing and EMM systems by prescribers and pharmacists and the factors influencing the acceptance of these systems in public health facilities of Dessie city administration, North East Ethiopia using a modified Unified Theory of Acceptance and Use of Technology (UTAUT) model.

## Methods

### Study Area

The study was conducted across all public health facilities of Dessie city administration, Northeast Ethiopia. Dessie serves as the capital city of South Wollo Zone, located 401 km away from Addis Ababa, the nation’s capital, and stands as one of the most populated towns in the Amhara regional state. According to the 2007 population and housing census report, Dessie had an estimated total population of 151,094 individuals. The urban inhabitants comprised 67.5% of the total population, with females accounting for 51.7% of the total population (37).

Within the Dessie City administration, there are two public hospitals, namely Dessie Comprehensive Specialized Hospital and Boru Meda General Hospital, along with four private hospitals, and eight public health centers: Bwanbwa wuha, Dessie, Segno Gebya, Gerado, Metero, Tita, Kurkur and Boru silase health centers Additionally, there are over 25 private clinics serving the area. Together, these health facilities provide services to a catchment population exceeding 8 million people, encompassing regions such as Tigray and Afar.

### Study design and period

A cross-sectional survey was conducted in public health facilities of Dessie city administration from April 20 to June 25, 2022.

### Source and study population

All prescribers and pharmacists working at public health facilities in Dessie town were the source population for this study. The study population was all prescribers and pharmacists working at public health facilities in Dessie City who were willing to participate during the study period.

### Inclusion and exclusion criteria

Prescribers and pharmacists with at least six months of experience who were willing to participate in the study were included in the study.

Prescribers and pharmacists who were on annual or sick leave during the study period were also excluded from the study.

### Sample size determination

All physicians, health officers, and pharmacists who fulfilled the inclusion criteria were included in the study. During the survey time, there were a total of 99 physicians and 33 pharmacists working in Dessie Comprehensive Specialized Hospital. Additionally, there were 24 physicians and 13 pharmacists in Boru Meda Hospital. In public health centers, there were 27 and 5 health officers and pharmacists. Overall, among prescribers and pharmacists working in public health facilities, 201 participants fulfilled the inclusion criteria and were included in the study.

### Study variables

#### Dependent variable

The dependent variable in this study is the acceptance of e-prescribing and EMM systems.

#### Independent variables

The independent variables in this study were performance expectancy, effort expectancy, perceived credibility, social influence, price value, and facilitating conditions.

#### Guiding model

The UTAUT model aims to integrate individual technology acceptance theories into a unified theoretical model, encompassing key elements from eight established models: Theory of Reasoned Action (TRA), Technology Acceptance Model (TAM), Motivational Model (MM), Theory of Planned Behavior (TPB), Diffusion of Innovation (DoI), Social Cognitive Theory (SCT), and model of personal computer use. The UTAUT has four key components: performance expectancy (PE), effort expectancy, social influence, and facilitating conditions. These components play a pivotal role in shaping individuals’ intentions to adopt information technology. Notably, UTAUT demonstrates a superior explanatory power, accounting for 70% of the variance in adoption behavior, compared to the earlier theories which typically explained only 30-40% of the variance (38).

This study hypothesized that the core constructs described in the UTAUT model are valid for measuring the behavioral intention of prescribers and pharmacists to use e-prescribing and EMM systems. Furthermore, this study identifies perceived credibility and price value as additional determinants of acceptance of technology. Therefore, the following hypotheses were defined. The study model is presented in Figure 1.

**Figure 1:**
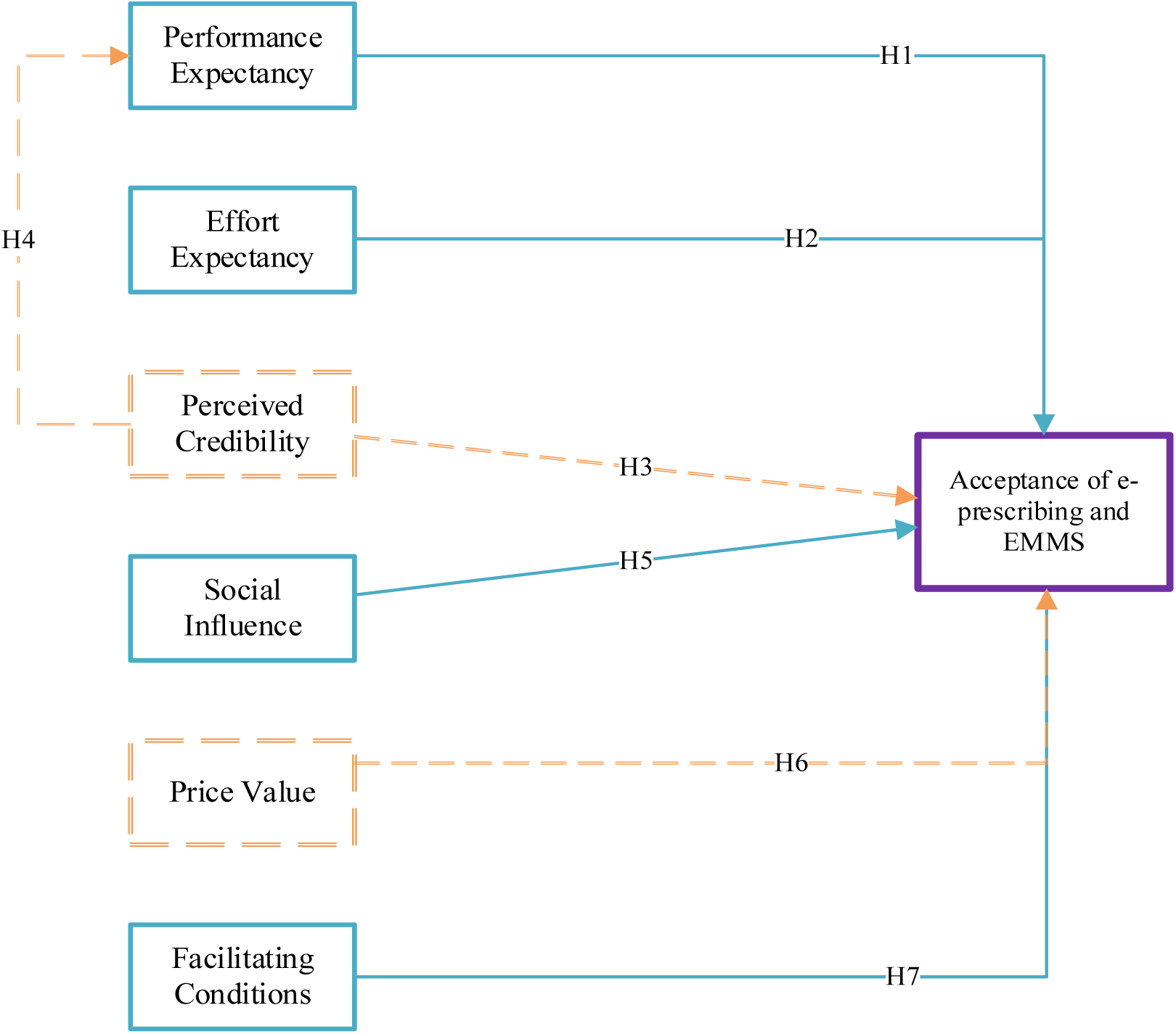
The research model - H: Hypothesis, Solid line: UTAUT model, Broken line: proposed modification
- H1: Performance expectancy has a positive impact on the acceptance of e-prescribing and EMM systems.
- H2: Effort expectancy has a positive impact on the acceptance of e-prescribing and EMM systems.
- H3: Perceived credibility (PC) on e-prescribing and EMM technologies has a positive impact on the acceptance of these systems.
- H4: Perceived credibility on e-prescribing and EMM technologies has a positive impact on performance expectancy.
- H5: Perceived social influence has a positive impact on the acceptance of e-prescribing and EMM systems.
- H6: Perceived price value has a positive impact on the acceptance of e-prescribing and EMM systems.
- H7: Perception of facilitating conditions has a positive impact on the acceptance of e-prescribing and EMM systems.

### Data collection and management

The data were collected by four trained pharmacists who had no working relation to the health facilities with the supervision of the investigators using a structured self-administered questionnaire. The questionnaire was based on the assumptions of the UTAUT model. This tool is validated and has been standardized through time, hence can be applied in different settings (39).

For the present study, 28 survey items were adapted from previous studies and modified to fit the context of e-prescribing and EMM systems. The questions for performance expectancy, effort expectancy, social influence, facilitating conditions, price value, and acceptance factors were adapted from Venkatesh et al (40), while those on the perceived credibility factor were adapted from Cohen et al (7). The questionnaire had two parts. The first part captured the sociodemographic characteristics of the study participants. The second part contained questions to assess the perceptions of prescribers and pharmacists on e-prescribing and EMM systems. Prescribers’ and pharmacists’ perceptions regarding e-prescribing and EMM systems were evaluated using a 5-point Likert-type scale. Scale scores were calculated by averaging the responses to the items within the scale. A mean score below 3 indicates a negative perception, while mean scores exceeding 3 denote a positive perception. Mean scores of 3 indicate a neutral perception. The questionnaire featured a concise explanation of e-prescribing and EMM systems to ensure participants’ familiarity and to mitigate inconsistencies in the interpretation of terms among them.

To ensure data quality, quality control checks were carried out before, during, and after data collection. The data collection instrument was pretested on 20 prescribers and pharmacists working in private health facilities. A one-day training was given to data collectors. All completed questionnaires were examined for completeness and consistency during data management, storage, and analysis.

### Data entry and processing

The completed questionnaires were individually coded and entered into a computer using Epi-info version 3.5.1, then exported to SPSS version 25.0 for analysis. Univariate analyses were used to describe the categorical variables, presenting frequency and percentage distributions of different characteristics. Means and standard deviations were used to describe continuous variables. The partial least squares approach to structural equation modeling was employed to test the hypothesized research model using SmartPLS version 4.0. The results are presented using figures and tables.

The constructs of the research were modeled with both reflective and formative indicators. Reflective constructs include performance expectancy, effort expectancy, social influence, perceived credibility, price value, and acceptance while facilitating conditions were modeled formatively. In partial least squares analysis, the evaluation of the outer model (measurement model) for reflective indicators is based on individual item reliability (indicator reliability), construct reliability, convergent validity, and discriminant validity (41). Individual item reliability (indicator reliability) was assessed through loadings, which are simple bivariate correlations of the indicators with their respective latent variable or construct (42). Construct reliability or internal consistency allows us to assess the extent to which indicators (observable variables) are measuring the latent variables. Composite reliability (ρc) and Cronbach’s α (42, 43) are commonly used for this purpose. However, composite reliability (CR) is more suitable for partial least squares analysis because it does not assume equal weighting for all indicators (44). The reliability of the measurement model was analyzed using Cronbach’s α, rho_A, and composite reliability.

Convergent validity of the measurement model was assessed using average variance extracted (AVE). An AVE of 0.50 or higher indicates that the construct explains more than half of the variance in its indicators (42). Discriminant validity, on the other hand, indicates the extent to which a given construct differs from other constructs (43). The discriminant validity of the model was assessed using the Fornell-Larcker Criterion (45) and the heterotrait-monotrait ratio of correlations (HTMT) (46).

Structural Models specify the relationships between the constructs (47) and reflect the hypothesized paths in the research framework (48). It was evaluated to test the hypotheses of the study. The strength of each causal relationship is measured using path coefficients, which indicate the causal effects of the independent variables on the dependent variable (49). The significance of the paths was determined by bootstrap resampling, which is utilized to generate standard errors for calculating t-values. A P-value less than 0.05 with a 95% confidence interval, was considered statistically significant.

### Operational definitions

Performance expectancy: the degree to which an individual believes using the e-prescribing and EMM systems will result in better job performance (38).

Effort expectancy: the degree to which an individual perceives using the e-prescribing and EMM systems is free of effort (38).

Perceived credibility: refers to an individual’s trust or confidence in the e-prescribing and EMM systems (50).

Social influence: is the degree to which an individual perceives that important others believe he or she should use e-prescribing and EMM systems (38).

Price value: is an individual’s evaluation of the perceived benefits of the e-prescribing and EMM systems and the monetary cost of using these systems (40).

Facilitating conditions: are defined as the degree to which an individual believes that there exist organizational and technical infrastructures that support the use of e-prescribing and EMM systems (38).

## Results

### Psychometric properties of the model

The measurement model was assessed for the reliability and validity of the models’ constructs (supplementary table A1). In this study, all items in the model exhibited loadings greater than 0.5, which is the minimum acceptable value (51). While factor loadings greater than 0.7 are desirable (52), items with outer loadings ranging from 0.40 to 0.708 should only be considered for removal if their deletion results in an increase in AVE or CR above the minimum acceptable value (53).

In the study, the removal of the items (EE4, loading = 0.696 and EE6, loading = 0.582) would not significantly impact the reliability of the model because the AVE and CR values for the construct effort expectancy already surpass the threshold values.

Statistics for both Cronbach’s α and CR exceeded the recommended value of 0.70 (54) for all the constructs. Additionally, the rho_A values extracted were also above 0.70(47) and all the rho_A values fell between Cronbach’s α and CR scores (55).

Convergent validity of the reflective measurement model was established, as indicated by AVE scores exceeding for all the constructs. Discriminant validity, which suggests that the square root of the AVE of each latent variable should be greater than its correlations with any other latent variables in the model (45), was confirmed. This condition is satisfied for all latent variables in the model (Table 1).

**Table 1:** Fornell-Larcker Criterion.

|  | **A** | **EE** | **FC** | **PC** | **PE** | **PV** | **SI** |
| --- | --- | --- | --- | --- | --- | --- | --- |
| *A* | 0.848* |  |  |  |  |  |  |
| *EE* | 0.51 | 0.72* |  |  |  |  |  |
| *FC* | 0.602 | 0.406 | ** |  |  |  |  |
| *PC* | 0.451 | 0.366 | 0.339 | 0.814* |  |  |  |
| *PE* | 0.59 | 0.37 | 0.394 | 0.359 | 0.818* |  |  |
| *PV* | 0.627 | 0.386 | 0.48 | 0.47 | 0.553 | 0.834* |  |
| *SI* | 0.52 | 0.344 | 0.592 | 0.342 | 0.482 | 0.488 | 0.83* |

\* The square root of the AVE of each latent variable
\*\* Not available as FC is a formative measurement model

Moreover, the HTMT ratio of correlations (46) was assessed and all the values were below the conservative threshold of 0.85 (supplementary table A2). Thus, discriminant validity was established. Regarding the items FC2 and FC3, although they exhibited nonsignificant outer weights (*p* values = .386 and .20), their outer loading exceeded 0.5 (loadings = 0.693 and 0.777). Therefore, the indicators (FC2 and FC3) were retained (53). Furthermore, analysis of collinearity within the measurement model showed that all the indicators had a variance inflation factor (VIF) below the cut-point of 5 (53). Hence, no items were removed from further analysis in the study.

### Socio-demographic characteristics

A total of 173 prescribers and pharmacists responded to the survey, after removing responses with significant missing data, 167 usable responses were extracted, resulting in a response rate of 83.08%. The majority of the respondents (72.5%) were males, and 73.7% were within the age group of 25 - 35 years. Nearly half (49.7%) of the respondents were general practitioners. The work experience of the study participants ranges from 6 months to 28 years with a modal class of 5 - 9.9 years. The majority of the respondents (71.3%) reported being able to perform basic computer tasks, while 8.4% had never used a computer before. The maximum number of daily prescriptions processed was 75, with a mean of 24.9 (Table 2).

**Table 2:** Socio-Demographic Characteristics of Prescribers and Pharmacists at Public Health Facilities of Dessie, Ethiopia, 2022 (N=167)

| Variables | Category | Frequency (%) |
| --- | --- | --- |
| Age group | Under 25 | 1 (0.6) |
|  | 25 – 34.99 | 123 (73.7) |
|  | 35 – 44.99 | 38 (22.8) |
|  | 45 – 54.99 | 4 (2.4) |
|  | 55 and above | 1 (0.6) |
| Sex | Male | 121 (72.5) |
|  | Female | 46 (27.5) |
| Marital Status | Single | 80 (47.9) |
|  | Married | 84 (50.3) |
|  | Divorced | 3 (1.8) |
| Profession | Physician | 101 (60.5%) |
|  | Health officer | 24 (14.4) |
|  | Pharmacist | 42 (25.1) |
| Work Experience (years) | below 5 | 74 (44.3) |
|  | 5 – 9.9 | 67 (40.1) |
|  | 10 – 14.9 | 16 (9.6) |
|  | 15 – 19.9 | 6 (3.6) |
|  | 20 and above | 4 (2.4) |
| Daily Prescription Volume | below 10 | 12 (7.2) |
|  | 10 – 19 | 32 (19.2) |
|  | 20 – 29 | 65 (38.9) |
|  | 30 - 39 | 35 (21.0) |
|  | 40 and above | 23 (13.8) |
| Computer Literacy | Never used a computer | 14 (8.4) |
|  | Can operate basic tasks using a computer | 119 (71.3) |
|  | Can operate advanced tasks using a computer | 34 (20.4) |

### Prescribers’ and pharmacists’ performance expectancy regarding e-prescribing and EMM system

More than half (58.1%) of the participants strongly agreed that using e-prescribing and EMM systems in their jobs would increase their productivity, while only 3% of them strongly disagreed. Additionally, about 43.7% of the participants agreed that using e-prescribing and EMM systems would make it easier to do their job. The mean performance expectancy was found to be 4.29±0.912 (Table 3).

**Table 3:** Prescribers’ and pharmacists’ performance expectancy regarding e-prescribing and e-MMSs, public health facilities of Dessie, 2022 (N=167)

|  | **Performance Expectancy (PE)** | **SD (%)** | **DA (%)** | **N (%)** | **A (%)** | **SA (%)** | **Mean (SD)** |
| --- | --- | --- | --- | --- | --- | --- | --- |
| 1 | Using e-prescribing and e-MM systems would improve my job Performance. | 5(3) | 2(1.2) | 19(11.4) | 67(40.1) | 74(44.3) | 4.22(0.913) |
| 2 | Using e-prescribing and e-MM systems in my job would enable me to accomplish tasks more quickly. | 5(3) | 7(4.2) | 13(7.8) | 58(34.7) | 84(49.7) | 4.25(0.98) |
| 3 | Using e-prescribing and e-MM systems would enhance my effectiveness on the job. | 4(2.4) | 3(1.8) | 14(8.4) | 54(32.3) | 92(55.1) | 4.36(0.893) |
| 4 | Using e-prescribing and e-MM systems in my job would increase my productivity. | 5(3) | 2(1.2) | 13(7.8) | 49(29.3) | 98(58.7) | 4.4(0.911) |
| 5 | Using e-prescribing and e-MM systems would make it easier to do my job. | 4(2.4) | 6(3.6) | 7(4.2) | 73(43.7) | 77(46.1) | 4.28(0.889) |
| 6 | I would find e-prescribing and e-MM systems useful in my job. | 4(2.4) | 3(1.8) | 15(9) | 65(38.9) | 80(47.9) | 4.28(0.884) |
| **Average** |  |  |  |  |  |  | **4.29(0.912)** |

*SD = strongly disagree, DA = Disagree, N = Neutral, SA = Strongly agree*

### Prescribers’ and pharmacists’ effort expectancy regarding e-prescribing and EMM system

Nearly half (48.1%) of the respondents strongly agreed that they would find it easy to get e-prescribing and EMM systems to do what they want them to do. About half (51.9%) of the prescribers and pharmacists strongly agreed that it would be easy for them to become skillful at using e-prescribing and EMM systems. The mean effort expectancy was found to be 4.26±0.87 (Table 4).

**Table 4:** Prescribers’ and pharmacists’ effort expectancy on e-prescribing and e-MMSs, public health facilities of Dessie, 2022 (N=167)

| Effort Expectancy (EE) | SD (%) | DA (%) | N (%) | A (%) | SA (%) | Mean (SD) |
| --- | --- | --- | --- | --- | --- | --- |
| 1. Learning to operate e-prescribing and e-MM systems would be easy for me. | 3(1.8) | 4(2.4) | 22(13.2) | 58(34.7) | 80(47.9) | 4.25(0.902) |
| 2. I would find it easy to get e-prescribing and e-MM systems to do what I want it to do. | 1(0.6) | 9(5.4) | 13(7.8) | 64(38.3) | 80(48.1) | 4.28(0.869) |
| 3. I would understand how to interact with e-prescribing and e-MM systems. | 3(1.8) | 4(2.4) | 17(10.2) | 73(43.7) | 70(41.9) | 4.22(0.858) |
| 4. I would find e-prescribing and e-MM systems to be flexible. | 1(0.6) | 7(4.2) | 14(8.4) | 66(39.5) | 79(47.3) | 4.29(0.837) |
| 5. It would be easy for me to become skillful at using e-prescribing and e-MM systems. | 1(0.6) | 8(4.8) | 13(7.8) | 59(35.3) | 86(51.9) | 4.32(0.837) |
| 6. I would find e-prescribing and e-MM systems easy to use. | 1(0.6) | 7(4.2) | 22(13.2) | 62(37.1) | 75(44.9) | 4.22(0.872) |
| **Average** |  |  |  |  |  | **4.26(0.866)** |

*SD = strongly disagree, DA = Disagree, N = Neutral, SA = Strongly agree*

### Facilitating conditions for acceptance of e-prescribing and EMM system

Nearly half (47.9%) of the participants strongly agreed that a specific person (or group) would be available for assistance with system difficulties, while only 1.8% of them strongly disagreed. The mean facilitating condition score was found to be 4.18±0.978 (Table 5).

**Table 5:** Prescribers’ and pharmacists’ facilitating conditions for e-prescribing and EMM systems, public health facilities of Dessie, 2022 (N=167)

| Facilitating Conditions (FC) | SD (%) | DA (%) | N (%) | A (%) | SA (%) | Mean (SD) |
| --- | --- | --- | --- | --- | --- | --- |
| 1. e-prescribing and e-MM systems would be compatible with other systems I use. | 5(3) | 8(4.8) | 18(10.8) | 69(41.3) | 67(40.1) | 4.11(0.982) |
| 2. I would have the resources necessary to use e-prescribing and e-MM systems. | 6(3.6) | 5(3.0) | 19(11.4) | 58(34.7) | 79(47.3) | 4.19(1.00) |
| 3. I would have the knowledge necessary to use e-prescribing and e-MM systems. | 5(3) | 7(4.2) | 14(8.4) | 65(38.9) | 76(45.5) | 4.20(0.971) |
| 4. A specific person (or group) would be available for assistance with system difficulties. | 3(1.8) | 10(6) | 14(8.4) | 60(35.9) | 80(47.9) | 4.22(0.959) |
|  |  |  |  |  |  | **Average 4.18(0.978)** |

*SD = strongly disagree, DA = Disagree, N = Neutral, SA = Strongly agree*

### Social influence on the acceptance of e-prescribing and EMM system by prescribers and pharmacists

Nearly half (47.9%) of the participants strongly agreed that people who influence their behavior and whose opinions they value think they should use e-prescribing and EMM systems. The mean social influence score was found to be 4.24±0.895 (Table 6).

**Table 6:** Prescribers’ and pharmacists’ social influences on e-prescribing and e-MMSs, public health facilities of Dessie, 2022 (N=167)

| Social Influence (SI) | SD (%) | DA (%) | N (%) | A (%) | SA (%) | Mean (SD) |
| --- | --- | --- | --- | --- | --- | --- |
| 1. People who are important to me think that I should use e-prescribing and e-MM systems. | 0 | 11(6.6) | 16(9.6) | 65(38.9) | 75(44.9) | 4.22(0.874) |
| 2. People who influence my behavior think that I should use e-prescribing and e-MM systems. | 3(1.8) | 9(5.4) | 14(8.4) | 61(36.5) | 80(47.9) | 4.23(0.944) |
| 3. People whose opinions I value think I should use e-prescribing and e-MM systems. | 2(1.2) | 5(3) | 19(11.4) | 61(36.5) | 80(47.9) | 4.27(0.867) |
| Average |  |  |  |  |  | 4.24(0.895) |

SD = strongly disagree, DA = Disagree, N = Neutral, SA = Strongly agree

### Prescribers’ and pharmacists’ perceived credibility of e-prescribing and EMM system

Half (50.3%) of the participants strongly agreed with the statement “I believe e-prescribing and EMM systems can be counted on to fulfill their function well”, while only 1.2% of them strongly disagreed with this statement. The mean perceived credibility of prescribers and pharmacists on e-prescribing and EMM systems was 4.26±0.854 (Table 7).

**Table 7:** Prescribers’ and pharmacists’ perceived credibility on e-prescribing and e-MMSs, public health facilities of Dessie, 2022 (N=167)

| Perceived Credibility (PC) | SD (%) | D (%) | N (%) | A (%) | SA (%) | Mean (SD) |
| --- | --- | --- | --- | --- | --- | --- |
| 1. I believe e-prescribing and e-MM systems will always meet my expectations. | 0 | 9(5.4) | 17(10.2) | 65(38.9) | 76(45.5) | 4.25(0.846) |
| 2. I believe e-prescribing and e-MM systems can be counted on to fulfil their function well. | 2(1.2) | 3(1.8) | 21(12.6) | 57(34.1) | 84(50.3) | 4.31(0.848) |
| 3. I believe e-prescribing and e-MM systems will be reliable. | 0 | 11(6.6) | 15(9) | 67(40.1) | 74(44.3) | 4.22(0.867) |
| **Average** |  |  |  |  |  | **4.26(0.854)** |

*SD = strongly disagree, DA = Disagree, N = Neutral, SA = Strongly agree*

### Prescribers’ and pharmacists’ price value regarding e-prescribing and EMM system

About half (51.5%) of the participants strongly agreed with the statement “at the current price, e-prescribing, and EMM systems provide good value”, while none of them strongly disagreed with this statement. The mean price value of e-prescribing and EMM systems was 4.26±0.892 (Table 8).

**Table 8:** Prescribers’ and pharmacists’ price value of e-prescribing and e-MMSs, public health facilities of Dessie, 2022 (N=167)

| Price Value (PV) | SD (%) | DA (%) | N (%) | A (%) | SA (%) | Mean (SD) |
| --- | --- | --- | --- | --- | --- | --- |
| 1. I believe e-prescribing and e-MM systems are reasonably priced. | 2(1.2) | 6(3.6) | 22(13.2) | 64(38.3) | 73 (43.7) | 4.20(0.887) |
| 2. I believe e-prescribing and e-MM systems have good value for money. | 4(2.4) | 4(2.4) | 20(12) | 56(33.5) | 83 (49.7) | 4.26(0.931) |
| 3. At the current price, e-prescribing and e-MM systems provide good value. | 0 | 10(6) | 13(7.8) | 58(34.7%) | 86 (51.5) | 4.32(0.858) |
| Average |  |  |  |  |  | 4.26(0.892) |

*SD = strongly disagree, DA = Disagree, N = Neutral, SA = Strongly agree*

### Prescribers’ and pharmacists’ acceptance of e-prescribing and EMM system

More than half (54.5%) of the participants strongly agreed with the statement “I plan to use e-prescribing and EMM systems, given the opportunity”. The mean acceptance of e-prescribing and EMMS by prescribers and pharmacists was 4.28± 0.963 (Table 9).

**Table 9:** Prescribers’ and pharmacists’ acceptance of e-prescribing and e-MMSs, public health facilities of Dessie city administration, 2022 (N=167)

| Acceptance (A) | SD (%) | DA (%) | N (%) | A (%) | SA (%) | Mean (SD) |
| --- | --- | --- | --- | --- | --- | --- |
| 1. I intend to use e-prescribing and e-MM systems, given the opportunity. | 5(3) | 5(3) | 6(3.6) | 75(44.9) | 76(45.5) | 4.27(0.910) |
| 2. I predict I would use e-prescribing and e-MM systems, given the opportunity. | 7(4.2) | 4(2.4) | 9(5.4) | 62(37.1) | 85(50.9) | 4.28(0.981) |
| 3. I plan to use e-prescribing and e-MM systems, given the opportunity. | 6(3.6) | 7(4.2) | 10(6) | 53(31.7) | 91(54.5) | 4.29(1.01) |
| **Average** |  |  |  |  |  | **4.28(0.963)** |

*SD = strongly disagree, DA = Disagree, N = Neutral, SA = Strongly agree*

### Structural model and hypotheses testing

The hypothesized structural model is presented in Figure 2. The model explains 60.2% of the variance in prescribers’ and pharmacists’ acceptance of e-prescribing and EMM systems. Out of the seven hypotheses proposed in this study, five were supported. The results of the analysis indicate that facilitating conditions exerted the most substantial influence on prescribers’ and pharmacists’ acceptance of e-prescribing and EMM systems. The results are presented in Table 10.

**Figure 2:**
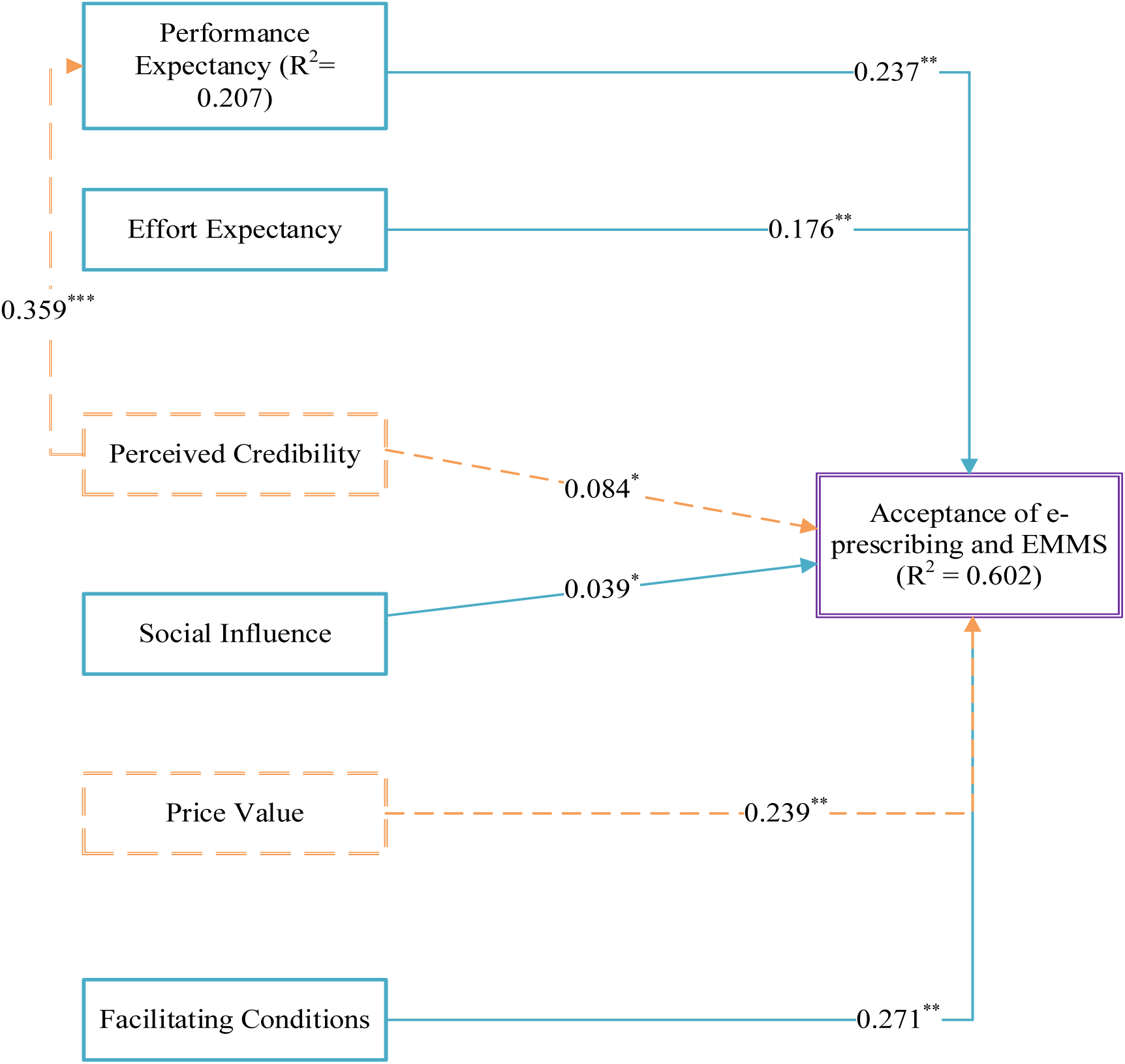
Partial least square test of the structural model *\*\*\* = p < .001; ** = p < .01; * = p > .05*

**Table 10:**
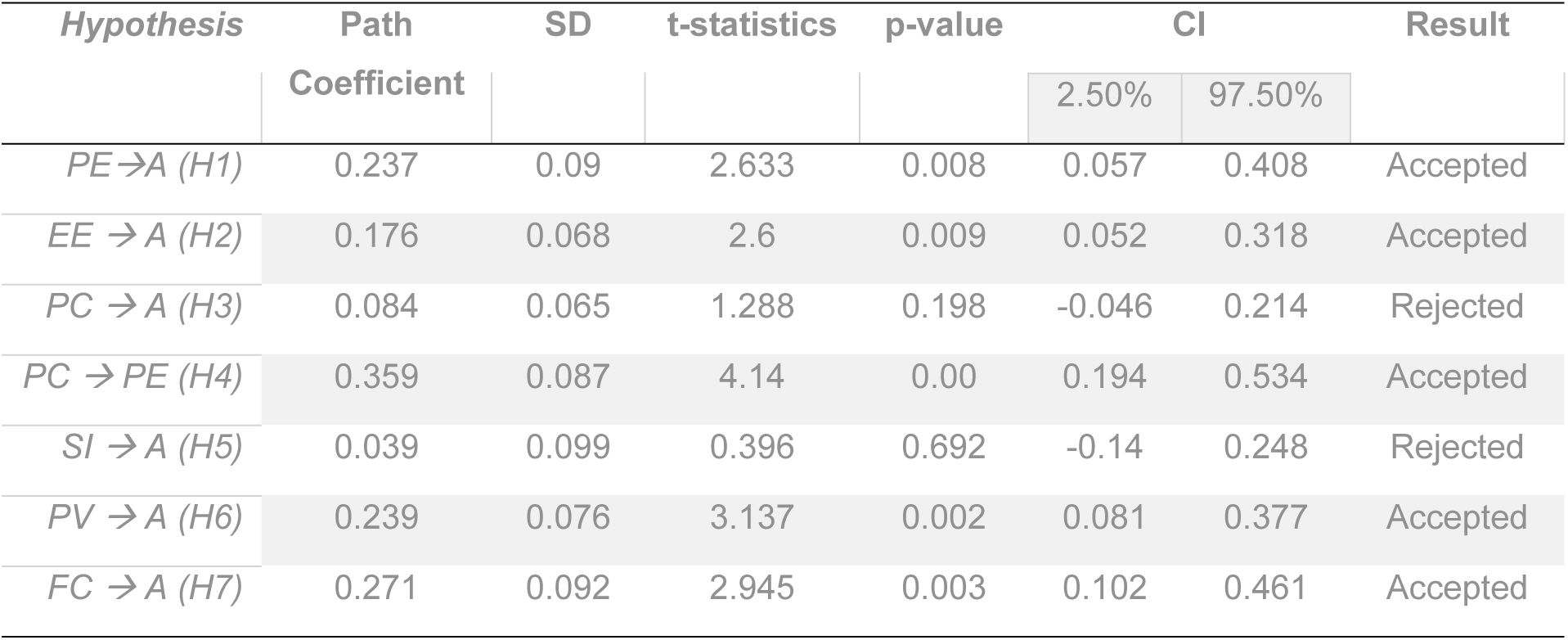
Regression coefficients from the structural model assessment and their corresponding results.

| *Hypothesis* | *Path* | *SD* | *t-statistics* | *p-value* | *CI* |  | *Result* |
| --- | --- | --- | --- | --- | --- | --- | --- |
|  | *Coefficient* |  |  |  | 2.50% | 97.50% |  |
| *PE → A (H1)* | 0.237 | 0.09 | 2.633 | 0.008 | 0.057 | 0.408 | Accepted |
| *EE → A (H2)* | 0.176 | 0.068 | 2.6 | 0.009 | 0.052 | 0.318 | Accepted |
| *PC → A (H3)* | 0.084 | 0.065 | 1.288 | 0.198 | -0.046 | 0.214 | Rejected |
| *PC → PE (H4)* | 0.359 | 0.087 | 4.14 | 0.00 | 0.194 | 0.534 | Accepted |
| *SI → A (H5)* | 0.039 | 0.099 | 0.396 | 0.692 | -0.14 | 0.248 | Rejected |
| *PV → A (H6)* | 0.239 | 0.076 | 3.137 | 0.002 | 0.081 | 0.377 | Accepted |
| *FC → A (H7)* | 0.271 | 0.092 | 2.945 | 0.003 | 0.102 | 0.461 | Accepted |

### H1: Performance expectancy has a positive impact on the acceptance of e-prescribing and EMM systems

A significant (p < 0.01) relationship was observed between performance expectancy and the acceptance of e-prescribing and EMM systems. Therefore, performance expectancy positively influences the acceptance of e-prescribing and EMM systems.

### H2: Effort expectancy has a positive impact on the acceptance of e-prescribing and EMM systems

The p-value associated with effort expectancy was found to be 0.009, indicating a significant relationship between effort expectancy and acceptance of e-prescribing and EMM systems. Hence, effort expectancy positively affects the acceptance of e-prescribing and E-MM systems.

### H3: Perceived credibility on e-prescribing and EMM technologies has a positive impact on the acceptance of these systems

The p-value associated with perceived credibility is 0.198, indicating that there is no significant direct relationship between perceived credibility and acceptance of e-prescribing and EMM systems.

### H4: Perceived credibility on e-prescribing and EMM technologies has a positive impact on performance expectancy

The results showed that there is a significant relationship between perceived credibility and performance expectancy (P = 0.00).

### H5: Perceived social influence has a positive impact on the acceptance of e-prescribing and EMM systems

The p-value associated with social influence on acceptance of e-prescribing and EMM systems is 0.629, suggesting a non-significant relationship.

### H6: Perceived price value has a positive impact on the acceptance of e-prescribing and EMM systems

The p-value of price value is 0.002, indicating a significant relationship between price value and acceptance of e-prescribing and EMM systems. Hence, price value positively influences the acceptance of e-prescribing and EMM systems.

### H7: Perception of facilitating conditions has a positive impact on the acceptance of e-prescribing and EMM systems

The p-value of facilitating conditions is 0.003, indicating a significant relationship between facilitating conditions and acceptance of e-prescribing and EMM systems. Therefore, facilitating conditions positively affect the acceptance of e-prescribing and EMM systems.

### Mediation analysis

Mediation analysis was performed to assess the mediating effect of performance expectancy on the relationship between perceived credibility and acceptance (A). The result showed that performance expectancy serves as a significant mediator in the relationship between perceived credibility and acceptance of e-prescribing and EMM systems. Specifically, while the direct effect (PC → A) of perceived credibility on acceptance was not significant (p = 0.198), both the indirect effect (PC → PE → A) and the total effect were significant, with p-values of 0.03 and 0.014 respectively (Table 11). This suggests that the impact of perceived credibility on the acceptance of e-prescribing and E-MMS is fully mediated through performance expectancy.

**Table 11:** Mediator role of performance expectancy on the relationship between perceived credibility and acceptance.

| Indirect effect ($PC \rightarrow PE \rightarrow A$) | | | | | | Direct effect ($PC \rightarrow A$) | | Total Effect ($PC \rightarrow A$) | |
| --- | --- | --- | --- | --- | --- | --- | --- | --- | --- |
| Path Coefficient<br>t | SD | t - statistics | p - value | CI |  | Path Coefficient | p -value | Path Coefficient | p -value |
|  |  |  |  | 2.50% | 97.50% |  |  |  |  |
| 0.085 | 0.039 | 2.165 | 0.03 | 0.017 | 0.168 | 0.084 | 0.198 | 0.169 | 0.014 |

## Discussion

This study aimed to assess prescribers’ and pharmacists’ perceptions towards the use of e-prescribing and EMM systems. It was found that factors such as performance expectancy, effort expectancy, price value, and facilitating conditions positively impact the acceptance of these systems. Interestingly, perceived credibility was not directly linked to acceptance; instead, its influence on acceptance was fully mediated by performance expectancy.

A recent meta-analysis revealed an overall prevalence of 57.6% of medication errors in Ethiopia (56). Additionally, it has been documented that the absence of e-health technologies such as e-prescribing systems is a significant contributing factor to the high rate of medication errors (56–59).In the present study, both prescribers and pharmacists believe that the adoption of e-prescribing and EMM systems would improve their job performance. In one study, the implementation of computerized order entry decision support resulted in a decrease in the rate of errors resulting in injury from 2.9 to 1.1 per 1000 patient days (15).

The current study revealed the significance of performance expectancy in driving the acceptance of e-prescribing and EMM systems. Healthcare professionals are more likely to embrace eHealth services when they anticipate tangible enhancements in clinical outcomes (**60**). Consequently, these systems must demonstrate clear performance improvements, which should be effectively communicated to clinicians. Moreover, the study findings indicate that performance expectancy acts as a mediator in the relationship between perceived credibility and acceptance of e-prescribing and EMM systems.

Previous studies have been inconclusive regarding factors that are most important to the acceptance of health information technology (7). However, in the present study, facilitating conditions are the most important factor in the acceptance of e-prescribing and EMM systems. It’s common for health facilities, particularly those in resource-limited settings, to lack the necessary infrastructures and skills to implement and operate new technologies. The findings of this study support the suggestion that vendor monitoring and outreach are essential to ensure healthcare practitioners have functional software and hardware (61). Therefore, these systems should be backed up by adequate resources, training programs, and technician support to enhance acceptance and implementation.

Various studies indicated that perceived ease of use is related to behavioral intention to use an information system (**7, 62, 63**). In this study, the mean effort expectancy was found to be 4.26, indicating that prescribers and pharmacists perceived that they would encounter few difficulties in using these systems once implemented. This result is higher than the reported in a study done in Kerala, India, (3.35) (64), which could be attributed to differences in the timing of studies. Notably, 88.6% of the participants in this study expressed a positive perception towards the use of e-prescribing and EMM systems. This finding is consistent with an internet-based survey conducted in the United States (83%) (65), suggesting that professionals who perceive these systems as easy to use are more likely to make an effort to try them.

Price value, which measures the health professionals’ cognitive tradeoff between perceived benefits and monetary cost associated with e-prescribing and EMM systems, has a positive relation with acceptance (β = 0.239, p = 0.002), which is consistent with the findings from a prior study (66). Healthcare professionals are inclined to adopt a system if they perceive the benefits of the system outweigh the costs of implementing and running the system.

One of the hypotheses of the present study was that social influence would positively influence the acceptance of e-prescribing and EMM systems. However, this hypothesis was not supported statistically (β = 0.039, p = 0.692), consistent with findings from previous studies (7, 67). This suggests that healthcare professionals may operate more autonomously and are less likely to be influenced by the opinions of influential individuals.

The findings of the present study revealed that the direct effect of perceived credibility on acceptance of e-prescribing and EMM systems was not significant (β = 0.084, p = 0.198), and this effect was fully mediated by performance expectancy, aligning with findings from a study conducted in South Africa (7).

## Limitations

The sampling frame of this study was limited to public health facilities Additionally, the moderating effects of sociodemographic characteristics and computer literacy on acceptance of e-prescribing and EMM systems were not assessed.

## Conclusions

Physicians and pharmacists had positive perceptions of the utilization of e-prescribing and EMM systems. Facilitating conditions, price value, performance expectancy, and effort expectancy (ease of use) were directly related to the acceptance of e-prescribing and EMM systems by prescribers and pharmacists. Perceived credibility had an indirect effect on the acceptance of e-prescribing and EMM systems, mediated by performance expectancy. However, its direct effect on acceptance was not significant.

Software vendors should prioritize the development of stable and trustworthy systems that are offered at a reasonable price. Additionally, health institutions should provide continuous technical support after the implementation of these systems to address any issues that may arise. Further research should be conducted to better understand the effects of sociodemographic characteristics and computer literacy on the acceptance of e-prescribing and EMM systems.

## Data Availability

The datasets used and/or analyzed during the current study are available from the corresponding author upon reasonable request.

## Abbreviations

A: Acceptance
AVE: Average Variance Extracted
CR: Composite Reliability
E-MM: Electronic medication management systems
e-Prescribing: Electronic prescribing
PE: Performance expectancy
PC: Perceived credibility
UTAUT: Unified Theory of Acceptance and Use of Technology

## Ethics approval and informed consent

The ethical approval was obtained from the College of Medicine and Health Science Research Ethics Review Committee (RERC), Wollo University (Ref. № - RCSPG-130-14). A letter of cooperation was written to Dessie Comprehensive Specialized Hospital, Boru Meda Hospital, and the health centers. Permission was obtained from the health facilities management. A written consent was also obtained from each study participant before commencing the study. Participant confidentiality was maintained by omitting names and other personal identifiers from the data collection tool. This study was conducted in accordance with the Declaration of Helsinki.

## Acknowledgments

The authors would like to express their gratitude to Wollo University for supporting materials and to Dessie Comprehensive Specialized Hospital, Boru Meda General Hospital, and the public health centers of Dessie city administration for allowing us to collect data. We are also grateful to data collectors and staff of both hospitals and health centers for their invaluable cooperation.

## Author contributions

All authors made substantial contributions to the conception and design, acquisition of data, or analysis and interpretation of data; took part in drafting the article or revising it critically for important intellectual content; agreed to submit to the current journal; gave final approval of the version to be published; and agree to be accountable for all aspects of the work.

## References

1. Eslami S, Abu-Hanna A, De Keizer NF. Evaluation of outpatient computerized physician medication order entry systems: a systematic review. Journal of the American Medical Informatics Association. 2007;14(4):400–6.

2. Donaldson MS, Corrigan JM, Kohn LT. To err is human: building a safer health system. 2000.

3. Čufar A, Droljc A, Orel A. Electronic medication ordering with integrated drug database and clinical decision support system. Quality of Life through Quality of Information: IOS Press; 2012. p. 693–7.

4. Westbrook JI, Reckmann M, Li L, Runciman WB, Burke R, Lo C, et al. Effects of two commercial electronic prescribing systems on prescribing error rates in hospital in-patients: a before and after study. PLoS medicine. 2012;9(1):e1001164.

5. Dovey S, Phillips R, Green L, Fryer G. Types of medical errors commonly reported by family physicians. American family physician. 2003;67(4):697-.

6. Velo GP, Minuz P. Medication errors: prescribing faults and prescription errors. British journal of clinical pharmacology. 2009;67(6):624–8.

7. Cohen JF, Bancilhon J-M, Jones M. South African physicians’ acceptance of e-prescribing technology: An empirical test of a modified UTAUT model. South African Computer Journal. 2013;50(1):43–54.

8. Bell DS, Cretin S, Marken RS, Landman AB. A conceptual framework for evaluating outpatient electronic prescribing systems based on their functional capabilities. Journal of the American Medical Informatics Association. 2004;11(1):60–70.

9. Schiff GD, Rucker TD. Computerized prescribing: building the electronic infrastructure for better medication usage. Jama. 1998;279(13):1024–9.

10. Shane R. Computerized physician order entry: challenges and opportunities. American journal of health-system pharmacy. 2002;59(3):286–8.

11. Tamblyn R, Huang A, Perreault R, Jacques A, Roy D, Hanley J, et al. The medical office of the 21st century (MOXXI): effectiveness of computerized decision-making support in reducing inappropriate prescribing in primary care. Cmaj. 2003;169(6):549–56.

12. Niazkhani Z, Pirnejad H, Berg M, Aarts J. The impact of computerized provider order entry systems on inpatient clinical workflow: a literature review. Journal of the American medical informatics Association. 2009;16(4):539–49.

13. Pearce R, Whyte I. Electronic medication management: is it a silver bullet? Australian prescriber. 2018;41(2):32.

14. Lindén-Lahti C, Kivivuori S-M, Lehtonen L, Schepel L, editors. Implementing a new electronic health record system in a university hospital: the effect on reported medication errors. Healthcare; 2022: MDPI.

15. Bates DW, Teich JM, Lee J, Seger D, Kuperman GJ, Ma’Luf N, et al. The impact of computerized physician order entry on medication error prevention. Journal of the American Medical Informatics Association. 1999;6(4):313–21.

16. Wolfstadt JI, Gurwitz JH, Field TS, Lee M, Kalkar S, Wu W, et al. The effect of computerized physician order entry with clinical decision support on the rates of adverse drug events: a systematic review. Journal of general internal medicine. 2008;23:451–8.

17. Beyene D, Abuye H, Tilahun G. Effect of auditable pharmaceutical services and transaction system on pharmaceutical service outcomes in public hospitals of SNNPR, Ethiopia. Integrated Pharmacy Research and Practice. 2020:185–94.

18. Davis FD. User acceptance of information technology: system characteristics, user perceptions and behavioral impacts. International journal of man-machine studies. 1993;38(3):475–87.

19. Pizzi LT, Suh D-C, Barone J, Nash DB. Factors related to physicians’ adoption of electronic prescribing: results from a national survey. American Journal of Medical Quality. 2005;20(1):22–32.

20. Anon. A Call to Action: Eliminate Handwritten Prescription Within 3 Years! : Institute for Safe Medication Practices, 2000; 2000.

21. Smithline N, Christenson E. Physicians and the Internet: understanding where we are and where we are going. The Journal of Ambulatory Care Management. 2001;24(4):39–53.

22. Thompson JN. Establishing guidelines for Internet-based prescribing. Southern medical journal. 2003;96(7):731.

23. Lo C, Burke R, Westbrook JI. Electronic medication management systems’ influence on hospital pharmacists’ work patterns. Journal of Pharmacy Practice and Research. 2010;40(2):106–10.

24. Hailiye Teferi G, Wonde TE, Tadele MM, Assaye BT, Hordofa ZR, Ahmed MH, et al. Perception of physicians towards electronic prescription system and associated factors at resource limited setting 2021: Cross sectional study. Plos one. 2022;17(3):e0262759.

25. Ketikidis P, Dimitrovski T, Lazuras L, Bath PA. Acceptance of health information technology in health professionals: An application of the revised technology acceptance model. Health informatics journal. 2012;18(2):124–34.

26. Ahmed MH, Bogale AD, Tilahun B, Kalayou MH, Klein J, Mengiste SA, et al. Intention to use electronic medical record and its predictors among health care providers at referral hospitals, north- West Ethiopia, 2019: using unified theory of acceptance and use technology 2 (UTAUT2) model. BMC Medical Informatics and Decision Making. 2020;20:1-11.

27. Masters K. Access to and use of the Internet by South African general practitioners. International journal of medical informatics. 2008;77(11):778–86.

28. Crow J, Broussard R, Dong L, Finn J, Wiley B, Geisler G, editors. A synthesis of research on ICT adoption and use by medical professionals in Sub-Saharan Africa. Proceedings of the 2nd ACM SIGHIT International Health Informatics Symposium; 2012.

29. Belle J-PV. Mobile technology adoption by doctors in public healthcare in the Western Cape, South Africa. 2006.

30. Banderker N, Van Belle J-P. Adoption of mobile technology by public healthcare doctors: A developing country perspective. International Journal of Healthcare Delivery Reform Initiatives (IJHDRI). 2009;1(3):38–54.

31. Banderker N, Van Belle J-P, editors. Mobile technology adoption by doctors in public healthcare in South Africa. Proceedings of the European Conference in Information Systems, Goteborg, Sweden; 2006.

32. West DM. Using mobile technology to improve maternal health and fight Ebola: A case study of mobile innovation in Nigeria. Center for Technological Innovation at Brookings. 2015;19:308–12.

33. Awol SM, Birhanu AY, Mekonnen ZA, Gashu KD, Shiferaw AM, Endehabtu BF, et al. Health professionals’ readiness and its associated factors to implement electronic medical record system in four selected primary hospitals in Ethiopia. Advances in medical education and practice. 2020:147–54.

34. Berihun B, Atnafu D, Sitotaw G. Willingness to Use Electronic Medical Record (EMR) System in Healthcare Facilities of Bahir Dar City, Northwest Ethiopia. BioMed Research International, 2020. 2020 (5): 2.

35. Biruk S, Yilma T, Andualem M, Tilahun B. Health Professionals’ readiness to implement electronic medical record system at three hospitals in Ethiopia: a cross sectional study. BMC medical informatics and decision making. 2014;14:1–8.

36. Yehualashet G, Asemahagn M, Tilahun B. The attitude towards and use of electronic medical record system by health professionals at a referral hospital in northern Ethiopia: Cross-sectional study. Journal of Health Informatics in Africa. 2015;3(1).

37. Population EOot, Commission HC. Summary and statistical report of the 2007 population and housing census: population size by age and sex: Federal Democratic Republic of Ethiopia, Population Census Commission; 2008.

38. Venkatesh V, Morris MG, Davis GB, Davis FD. User acceptance of information technology: Toward a unified view. MIS quarterly. 2003:425–78.

39. Wrzosek N, Zimmermann A, Balwicki Ł, editors. Doctors’ perceptions of e-prescribing upon its mandatory adoption in poland, using the unified theory of acceptance and use of technology method. Healthcare; 2020: MDPI.

40. Venkatesh V, Thong JY, Xu X. Consumer acceptance and use of information technology: extending the unified theory of acceptance and use of technology. MIS quarterly. 2012:157–78.

41. Hair JF, Sarstedt M, Ringle CM, Mena JA. An assessment of the use of partial least squares structural equation modeling in marketing research. Journal of the academy of marketing science. 2012;40:414–33.

42. Hair Jr JF, Sarstedt M, Hopkins L, Kuppelwieser VG. Partial least squares structural equation modeling (PLS-SEM): An emerging tool in business research. European business review. 2014;26(2):106–21.

43. Roldán JL, Sánchez-Franco MJ. Variance-based structural equation modeling: Guidelines for using partial least squares in information systems research. Research methodologies, innovations and philosophies in software systems engineering and information systems: IGI global; 2012. p. 193–221.

44. Chin WW. The partial least squares approach to structural equation modeling. Modern methods for business research. 1998;295(2):295–336.

45. Fornell C, Larcker DF. Structural equation models with unobservable variables and measurement error: Algebra and statistics. Sage publications Sage CA: Los Angeles, CA; 1981.

46. Henseler J, Ringle CM, Sarstedt M. A new criterion for assessing discriminant validity in variance-based structural equation modeling. Journal of the academy of marketing science. 2015;43:115–35.

47. Henseler J, Hubona G, Ray PA. Using PLS path modeling in new technology research: updated guidelines. Industrial management & data systems. 2016;116(1):2–20.

48. Latif KF, Sajjad A, Bashir R, Shaukat MB, Khan MB, Sahibzada UF. Revisiting the relationship between corporate social responsibility and organizational performance: The mediating role of team outcomes. Corporate Social Responsibility and Environmental Management. 2020;27(4):1630–41.

49. Lleras C. Path analysis. Encyclopedia of social measurement. 2005;3(1):25–30.

50. Khan IU, Yu Y, Hameed Z, Khan SU, Waheed A. Assessing the physicians’ acceptance of e-prescribing in a developing country: An extension of the UTAUT model with moderating effect of perceived organizational support. Journal of Global Information Management (JGIM). 2018;26(3):121–42.

51. Hair JF, Anderson RE, Babin BJ, Black WC. Multivariate data analysis: A global perspective (Vol. 7). Upper Saddle River, NJ: Pearson; 2010.

52. Vinzi VE, Chin WW, Henseler J, Wang H. Handbook of partial least squares: Springer; 2010.

53. Leguina A. A primer on partial least squares structural equation modeling (PLS-SEM). Taylor & Francis; 2015.

54. Wasko MM, Faraj S. Why should I share? Examining social capital and knowledge contribution in electronic networks of practice. MIS quarterly. 2005:35–57.

55. Sarstedt M, Ringle CM, Hair JF. Partial least squares structural equation modeling. Handbook of market research: Springer; 2021. p. 587–632.

56. Bifftu BB, Mekonnen BY. The magnitude of medication administration errors among nurses in Ethiopia: a systematic review and meta-analysis. Journal of Caring Sciences. 2020;9(1):1.

57. Agalu A, Ayele Y, Bedada W, Woldie M. Medication prescribing errors in the intensive care unit of Jimma University Specialized Hospital, Southwest Ethiopia. Journal of multidisciplinary healthcare. 2011:377–82.

58. 58. Feyissa D, Kebede B, Zewudie A, Mamo Y. Medication error and its contributing factors among pediatric patients diagnosed with infectious diseases admitted to Jimma University Medical Center, Southwest Ethiopia: prospective observational study. Integrated Pharmacy Research and Practice. 2020:147-53.

59. Sada O, Melkie A, Shibeshi W. Medication prescribing errors in the medical intensive care unit of Tikur Anbessa Specialized Hospital, Addis Ababa, Ethiopia. BMC research notes. 2015;8:1–7.

60. Nuq PA, Aubert B. Towards a better understanding of the intention to use eHealth services by medical professionals: the case of developing countries. International Journal of Healthcare Management. 2013;6(4):217–36.

61. Halamka J, Aranow M, Ascenzo C, Bates DW, Berry K, Debor G, et al. E-Prescribing collaboration in Massachusetts: early experiences from regional prescribing projects. Journal of the American Medical Informatics Association. 2006;13(3):239–44.

62. Aggelidis VP, Chatzoglou PD. Using a modified technology acceptance model in hospitals. International journal of medical informatics. 2009;78(2):115–26.

63. Chau PY, Hu PJ-H. Investigating healthcare professionals’ decisions to accept telemedicine technology: an empirical test of competing theories. Information & management. 2002;39(4):297–311.

64. Palappallil DS, Pinheiro C. Perceptions of prescribers towards electronic prescription: A pre-implementation evaluation. Journal of Young Pharmacists. 2018;10(3):313.

65. Jariwala KS, Holmes ER, Banahan III BF, McCaffrey III DJ. Adoption of and experience with e-prescribing by primary care physicians. Research in Social and Administrative Pharmacy. 2013;9(1):120–8.

66. Tavares J, Oliveira T. Electronic health record portal adoption: a cross country analysis. BMC medical informatics and decision making. 2017;17:1–17.

67. Duyck P, Pynoo B, Devolder P, Voet T, Adang L, Vercruysse J. User acceptance of a picture archiving and communication system. Methods of information in medicine. 2008;47(02):149–56.

